# Prevalence and Nature of Sexual Violence in Older Adults in Europe: A Critical Interpretive Synthesis of Evidence

**DOI:** 10.1101/2020.04.24.20077990

**Authors:** Anne Nobels, Christophe Vandeviver, Marie Beaulieu, Adina Cismaru Inescu, Laurent Nisen, Nele Van Den Noortgate, Tom Vander Beken, Gilbert Lemmens, Ines Keygnaert

**Author notes:** Direct correspondence to: Anne Nobels, International Centre for Reproductive Health- Ghent University, C. Heymanslaan 10 entrance 75- ICRH, 9000 Gent, Belgium.

## Abstract

Sexual violence (SV) is an important public health issue with a potential major impact on victims and their peers, offspring and community. However, SV in older adults is under-researched. This paper aims to establish the prevalence and nature of SV in older adults in Europe, link this with existing policies and health care workers’ response to sexual health needs in older age and critically revise the current used frameworks in public health research.

We applied a Critical Interpretative Synthesis. After the first phase of purposive sampling we included 14 references. Another 14 references were included after the second phase of theoretical sampling. We ultimately included 16 peer-reviewed articles and 12 documents from the grey literature.

0.0% to 3.1% of older adults in Europe were sexually victimised in the past year. Lifetime prevalence of SV was 6.3%. Information on specific risk factors and assailants committing SV in old age is non-existing. Although in theory policy makers increasingly recognise the importance of sexual health in older age, SV in older adults is not mentioned in policy documents on sexual and reproductive health and rights and ageing. In clinical practice, the sexual health needs of older adults remain often unmet. Knowledge about SV in older adults is still limited. Ongoing research does not fully grasp the complexity of SV in older adults. Greater awareness about this topic could contribute to a revision of current policies and health care practices, leading to more tailored care for older victims of SV.

## INTRODUCTION

Since the 1990s, sexual violence (SV) [1] has increasingly been considered a public health problem of major societal and judicial concern [2-4]. SV is defined by the World Health Organisation (WHO) as “Every sexual act directed against a person’s will, by any person regardless of their relationship to the victim, in any setting” [1]. In recent years, several European studies reported that around 30% of women and 10-27% of men experienced at least one incident of SV between 16 and 25 years [5-11].

In public health research, SV in older adults is defined in the broader context of elder abuse and neglect, which is considered as “A single or repeated act, or lack of appropriate action, occurring within any relationship where there is an expectation of trust which causes harm or distress to an older person’’ [4]. It comprises a broad range of abusive behaviours including psychological abuse, financial abuse, physical abuse, sexual abuse, and neglect.

Worldwide one in six older adults seems to be affected by elder abuse and neglect, and 0.9% of older adults was sexually victimised in the past year [12]. In older women past year SV prevalence rates increased to 2.2% [13]. Since 30% of the European population is estimated to be 65 years or older by Extensive research indicates that not only victims of SV, but also their peers, offspring, and community may endure long-lasting sexual, reproductive, physical, and mental health problems [15]. Moreover, child sexual abuse can result in several long term consequences that might persist through adulthood [16, 17] and presumably later life. Several studies show that older adults who experienced adverse childhood experiences (ACEs) (including child sexual abuse (CSA)) had an increased risk of both mental and physical health problems in later life [18-21] and that the negative effects of ACEs on the risk of various health problems are unaffected by social or secular changes [22].

Notwithstanding the fact that older adults are at risk of SV [23], manifestations of both short and long term consequences of SV are rarely recognised in older age [24]. Additionally, the sexual health of older adults is often ignored [25], although several studies have shown that sexuality remains important in older age [26-28]. Therefore, increased attention to older adults in research, policy and health care practices concerning sexual health and sexual violence is urgently demanded [29].

In recent years several reviews have been published on SV in older adults looking at it from a criminal perspective [30-32]. They focus on criminal cases and describe assailant characteristics, barriers to disclosure and justice response. However, research on SV in older adults from a public health perspective is lacking.

This review is the first to explore SV in older adults outside of the context of crime. We critically review the evidence on the prevalence and nature of SV in older adults in Europe and link this with existing policies on prevention and response to SV in older adults and health care workers’ response to sexual health needs of older adults. We reflect on the currently used frameworks in public health research on SV in older adults. Based on the existing evidence, we formulate recommendations for future research, policies and health care practices. 2060 [14], elder abuse, including SV, has to become an increasing public health concern [12].

## METHODS

### Critical Interpretive Synthesis

In order to appraise the evidence on SV in older adults, we conducted a Critical Interpretive Synthesis (CIS). The CIS method uses techniques from grounded theory and processes from systematic review [33]. It is designed to handle a heterogeneous sample of documents, both academic, as well as non-peer reviewed reports, policies and legal frameworks. Hence, a CIS offers authors the opportunity to include documents that would not meet the inclusion criteria for a systematic review. This leads to a broader scope, helping us to better understand the complexity of SV at older age. A CIS starts from an open research question, applying a dynamic and recursive approach, which allows us to identify questions during the review process [34]. The ultimate goal is the construction of new theoretical frameworks, that may serve as a starting point for future research [33].

### Sample of studies

During the CIS method we used a two-stage sample process: (1) purposive sampling and (2) theoretical sampling based on the principles of theoretical saturation [35].

After the first phase of purposive sampling we included 14 references. Another 14 references were included after the second phase of theoretical sampling based on the principles of theoretical saturation. We ultimately included 16 peer-reviewed articles and 12 documents from the grey literature (7 reports, 4 policy documents and 1 book chapter) in our review (Figure 1).

**Figure 1.**
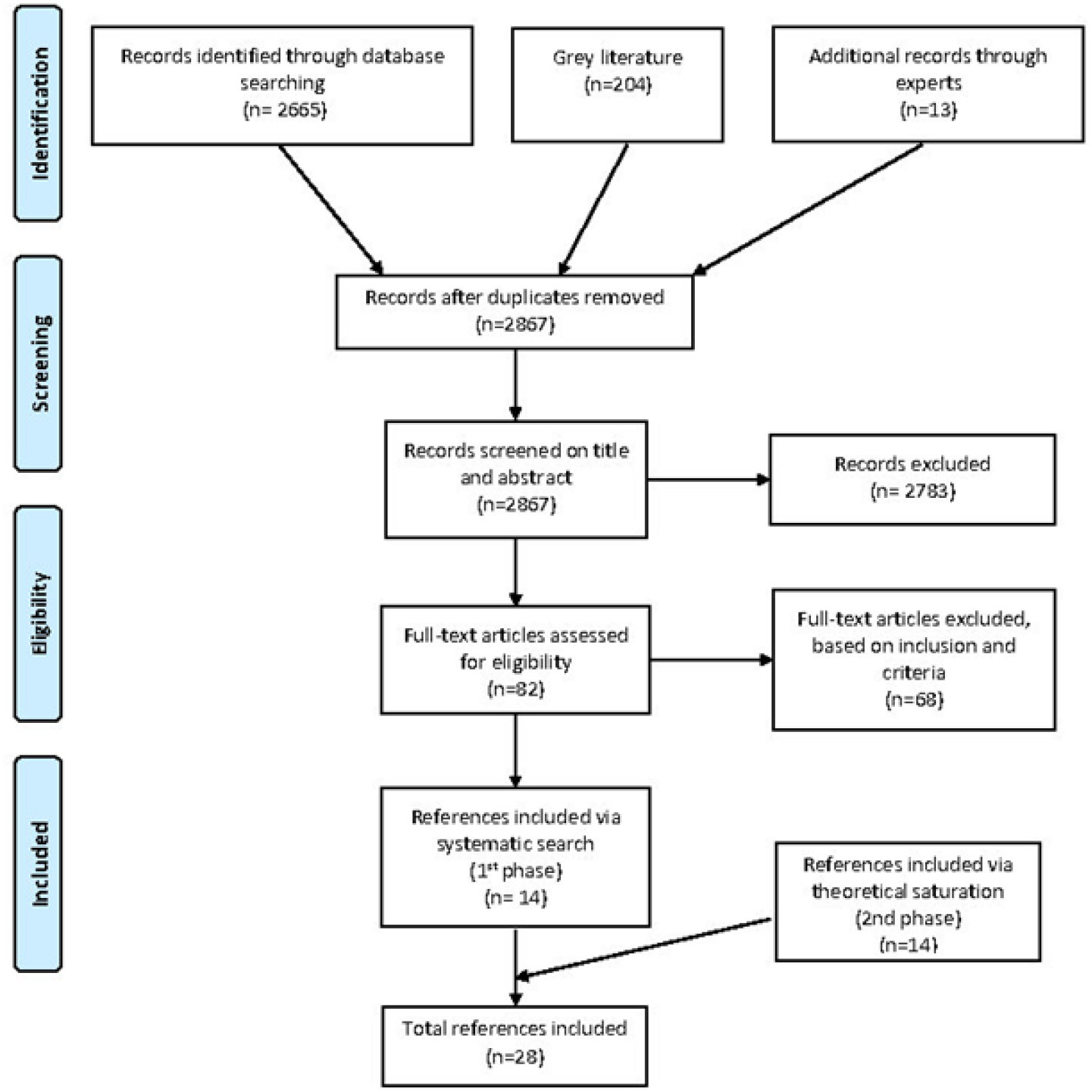
Flow diagram of reference selection process.

#### Phase 1: Purposive sampling

In the first phase of the purposive sampling we used a comprehensive four-step search strategy. First, we performed a systematic search in five databases: Pubmed, Web of Science, Google Scholar, Embase and Cochrane Central using following search terms: ‘sexual violence’, ‘sexual abuse’, ‘intimate partner violence’, ‘older adults’, ‘older people’, ‘elderly’, ‘Europe’, ‘prevalence’, elder abuse’, ‘elder abuse and neglect’ and ‘elder mistreatment’.

Second, reference lists of publications retrieved in the first step were screened for relevant studies. Third, additional web-based platforms including specialised journals, websites of notable international organizations (as United Nations (UN) and affiliated agencies, WHO), and NGOs working on elder abuse were searched and Google searches were reviewed for grey literature.

Finally, we contacted 14 national and international experts in the field of SV and elder abuse and neglect by email to provide further review to identify any studies that were missing up to November 30, 2019.

Literature was screened by two independent researchers. Peer-reviewed articles and grey documents were selected based on the following inclusion and exclusion criteria:

1. Literature published between January 2002 and November 2019 was included. In 2002 the UN called upon elimination of all forms of elder abuse and neglect in their Action Plan on Ageing [36]. After expert consultation, two studies conducted before 2002 were purposely included, because of their importance to the Belgian context [37, 38].
2. Only studies conducted in the WHO European region were included.
3. Older adults needed to be defined as a separate (research) population regardless of the cut-off age used for old age.
4. Only studies reporting on the self-reported prevalence rates and associated risk factors of sexual victimisation in older adults were included. We excluded studies using reports of family members or health care workers or studies using crime statistics.
5. In order to provide a comprehensive overview of victimisation rates in older adults, only literature focussing on community-dwelling older adults and older adults living in nursing homes were included.
6. Due to restrictions in the author’s language proficiency, only literature in English, Dutch and French was included.
7. Reviews of literature, methodological papers, commentaries and conference abstracts were excluded.

#### Phase 2: Theoretical sampling

In the second phase of theoretical sampling we researched existing policies on SV and elder abuse and neglect and policies on sexual health looking for specific prevention and response strategies concerning SV at old age. Additionally, we looked into different ways in which health care workers deal with sexual health needs, including sexual violence, in older age.

## RESULTS

In the results section we first discuss the results of the purposing sampling and then reflect on the additional literature retrieved from the theoretical sampling.

### Systematic review of prevalence and nature of SV at older age

#### Description of included studies (Table 1)

The 14 references selected for the review included information from 15 different countries: three studies from Belgium [37-39] one from Ireland [40], one from Israel [41], one from Italy [42], one from Poland [43], one from Portugal [44], three from Turkey [45-47], one from the United Kingdom [48] and two European multi-country studies [49, 50]. Twelve studies reported on community-dwelling older adults [37, 39-41, 43-50], one on older adults living in nursing homes [38] and one study comprised a mixed sample [42]. Moreover, nine studies were based on random samples [37, 39-41, 44, 47-50], five were convenience samples [38, 42, 43, 45, 46]. Five of the random samples were nationally-representative [37, 40, 41, 44, 48]. Five studies used the age of 60 as cut-off age for inclusion of older adults [42, 44, 46, 49, 50], five studies used 65 [37, 40, 41, 45, 47], one study 66 and one study used 70 [39]. Two studies did not report a cut-off age [38, 43]. Two studies only included older women [38, 50], one study included a majority of males [47], eight studies included a majority of female participants [37, 39-42, 44, 45, 49]. Three studies did not report a female-male ratio [43, 46, 48]. All studies, except two [42, 44], used face-to-face interviews. Participation rates ranged from 26 to 93%. One study reported on the last 3-months prevalence of SV [41], one on the last 6- months prevalence [43], eight on the last year prevalence [40, 42, 44-46, 48-50], one on the lifetime prevalence [37] and four on the prevalence from their cut-off age, being 60, 65 or 70 years old [37, 39, 40, 48]. One study in a nursing home population reported on the time period the older adults had resided in the nursing home [38]. One study did not report a time frame [47].

**Table 1.**
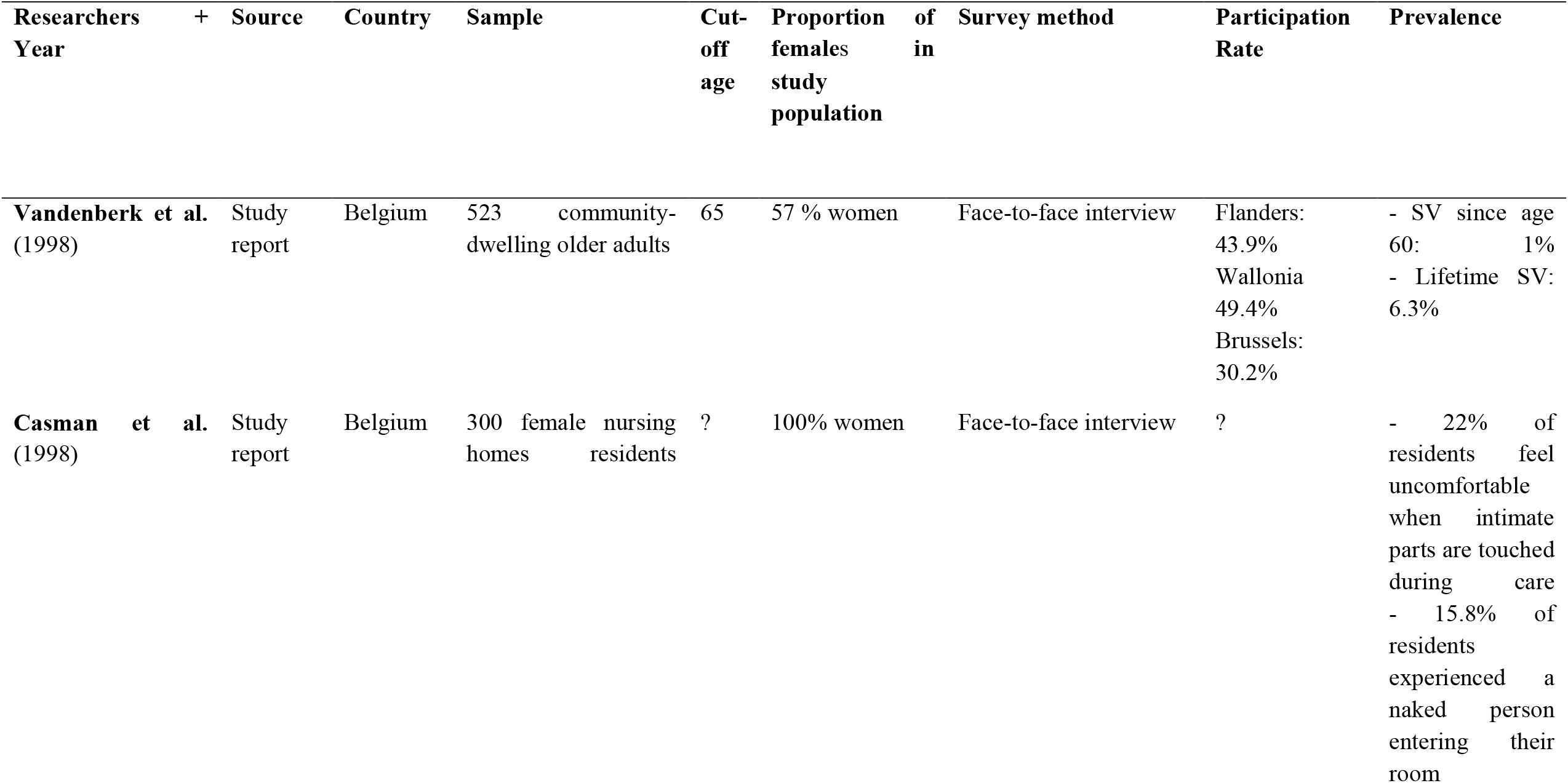
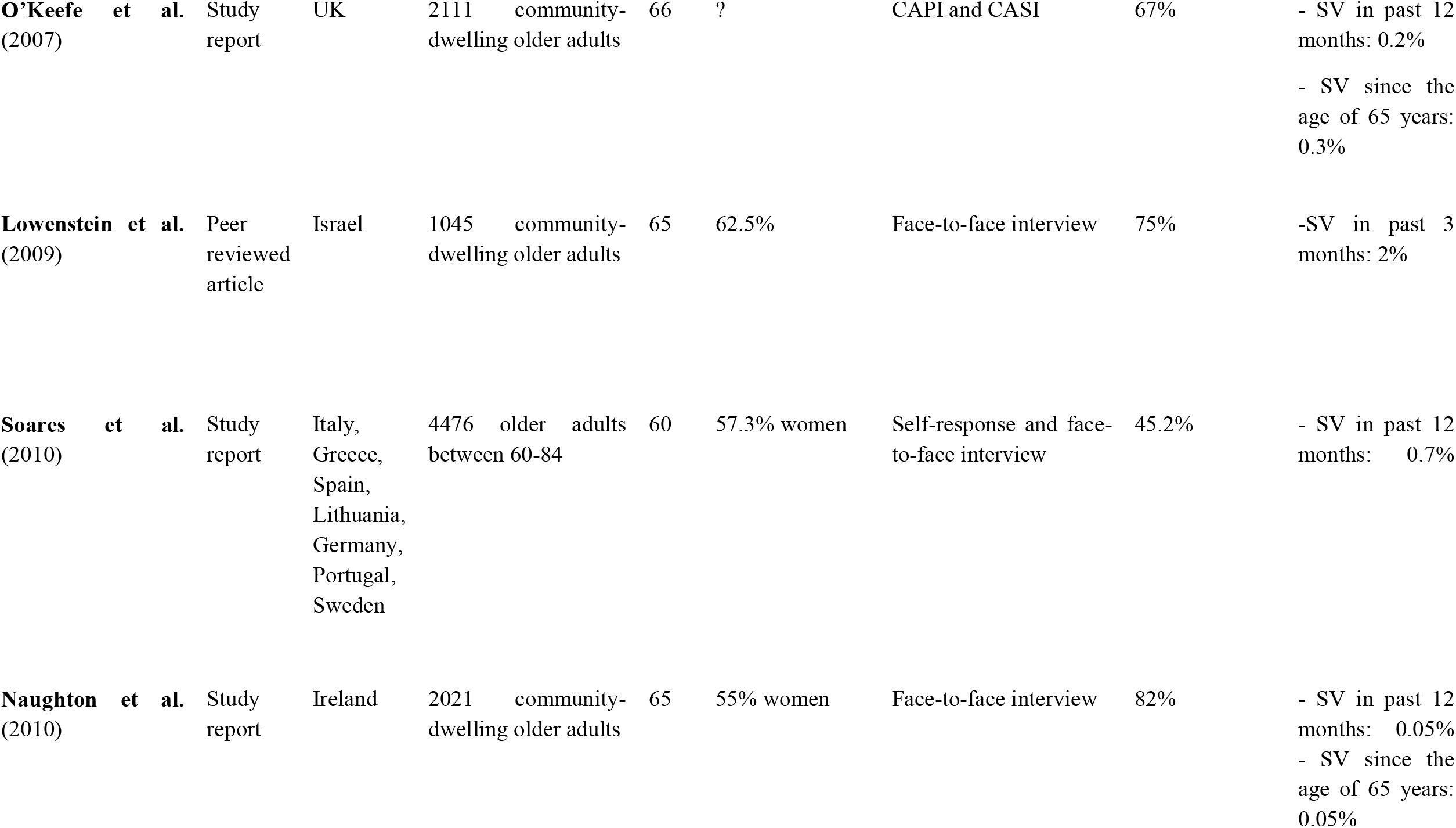
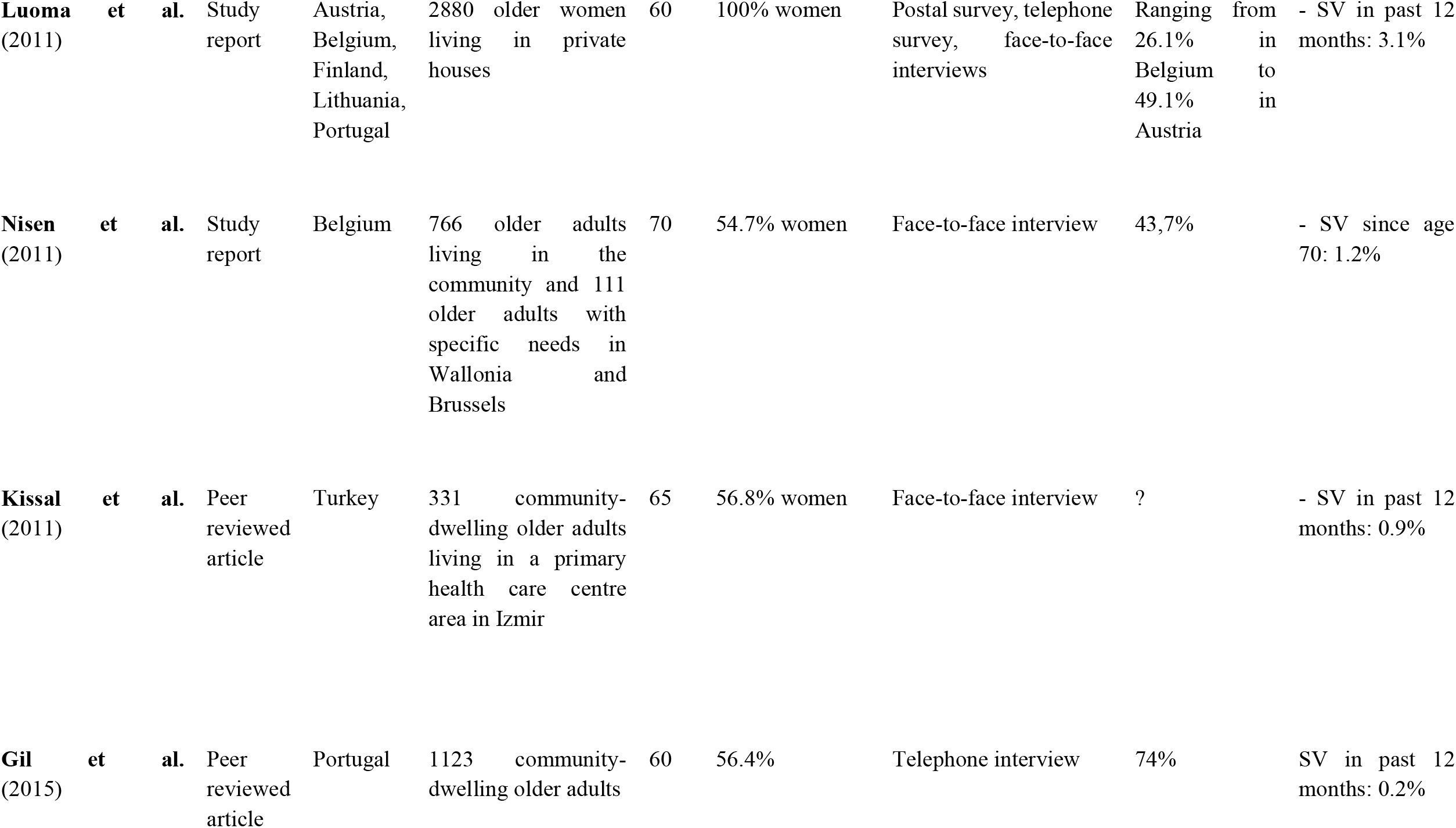
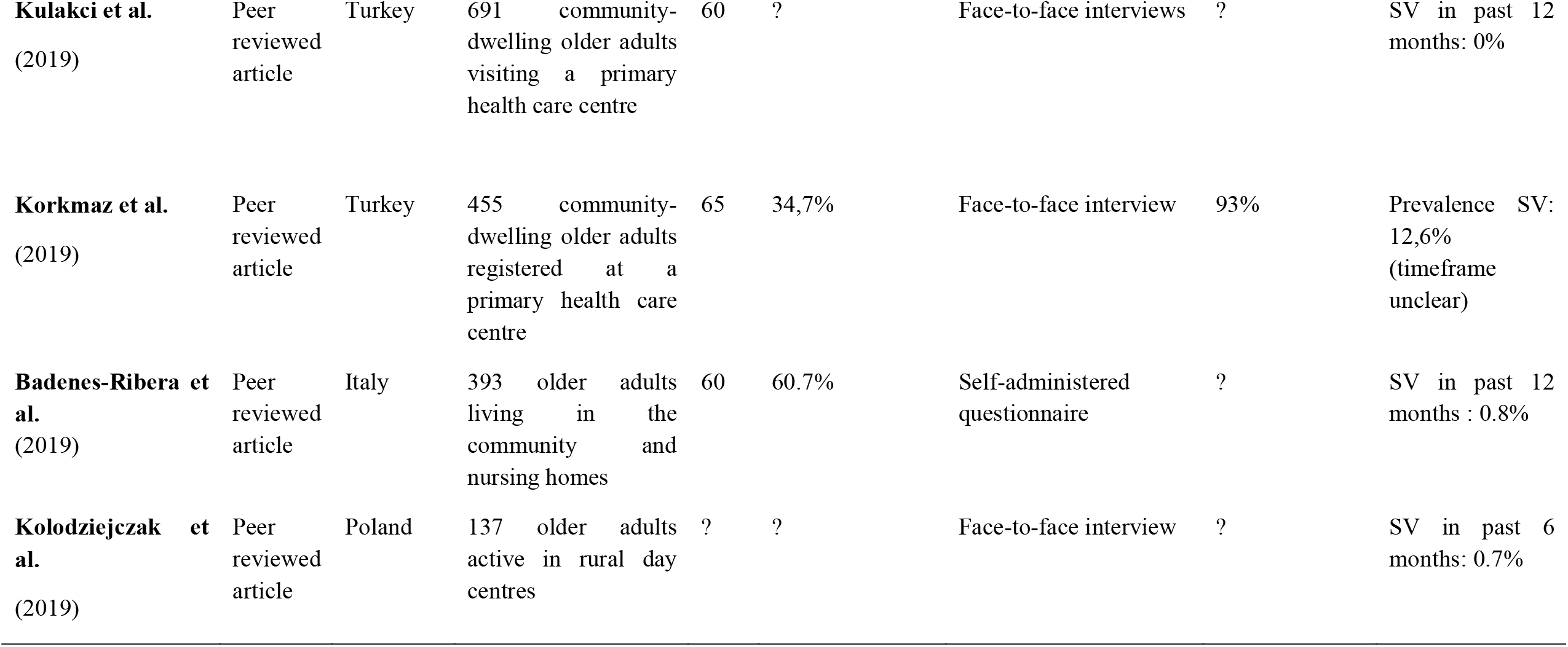
Description of included studies in phase 1. SV: Sexual Violence, CAPI: Computer assisted personal interviewing, CASI: Computer assisted self-interviewing

#### Prevalence of SV in older adults

In community-dwelling older adults the last year prevalence of SV varied between 0% to 3.1% [37, 40, 41, 44-46, 48, 50]. One study reporting on a mixed sample of 393 community-dwelling older adults and nursing home residents found a last-year prevalence of SV of 0.8% [42]. SV since the start of older age was reported by 0.05% to 1.2% of community-dwelling older adults [37, 39, 40]. One study reported a SV lifetime prevalence rate in community-dwelling older adults of 6.3% [37]. A study describing a population of 455 community-dwelling older adults registered at a primary health care centre in Turkey, found a 12.6% prevalence rate for SV. However, the time frame of this study was not specified [47].

The study on nursing home residents did not report the overall prevalence of SV, but the results gave some insight in to the magnitude of the problem in nursing homes. The study revealed that 22% of residents felt uncomfortable when their intimate parts were touched during care and 15.8% had experienced (directly or indirectly) a naked person entering their room [38].

#### Risk factors for SV at older age

None of the 14 studies included a separate analysis of the specific risk factors for SV in older adults. In the following paragraph we describe the risk factors for elder abuse and neglect, however a generalisation to the context of SV must be done with caution.

Concerning risk factors for elder abuse and neglect, several studies on community-dwelling older adults reported that female older adults and older adults with a prior history of abuse were at higher risk of being victimised [37, 40, 44, 45]. Other studies link a poor (perceived) health status and limited autonomy with a higher risk of being abused [38, 42, 47]. Abused older adults reported more physical and mental health problems, had lower social support and indicated more financial strain compared to non-abused older adults [39, 40, 44-47, 50]. Two Turkish studies showed a higher risk of abuse for illiterate older adults [46, 47].

Several studies indicated that the risk of being abused increased with age [40, 44, 47]. Concerning living conditions, two studies described a higher risk of abuse when living alone [44, 49] while others demonstrated that living with a partner or children increased the risk of abuse [45, 50].

#### Assailants of SV in older age

Six of the included studies reported on assailants who committed interpersonal abuse, which includes sexual, physical and psychological abuse, but excludes financial abuse and neglect [40-42, 44, 48, 50]. In five of these studies the most common assailants were the partner of the victim or other relatives and friends [40, 41, 44, 48, 50]. One study mentioned paid care givers as most commonly responsible for physical/sexual abuse [42]. One study revealed that the majority of assailants suffered from physical, mental and/or cognitive problems [41], another study found that 28% of the assailants had relationship problems [40]. Whereas assailants committing financial abuse were more likely to be young and live elsewhere, assailants performing interpersonal abuse were more likely to be male, older and living with the victim [40, 48].

One study reported separately on assailants who committed sexual abuse. They found that 30% of sexual abuse was perpetrated by friends, acquaintances and neighbours, 27% by ‘others’, 24% by partners, 3% by offspring and 3% other relatives [49].

### Policies on prevention and response to SV in older adults

Since the International Conference on Population and Development (ICPD) in Cairo in 1994 numerous international agreements have affirmed a commitment to universal access to sexual and reproductive health (SRH). At the ICPD25 summit in Nairobi in November 2019 this commitment was confirmed, and special attention was given to a comprehensive life course approach to sexual and reproductive health and rights. This life course approach recognises that people have different and changing sexual and reproductive health needs throughout their lives. In addition, it takes account of how sexual and reproductive health needs and decisions at one stage in life have implications for sexual and reproductive health outcomes and needs later in life [51].

In the WHO European Region, both the “Strategy Plan for Healthy Ageing in Europe 2012-2020” and the “Action Plan for Sexual and Reproductive Health: towards achieving the 2030 Agenda for Sustainable Development in Europe-leaving no one behind” called for more actions in improving the health of older people, including sexual health [52, 53]. Additionally, the WHO Global “Strategy and Action Plan on Ageing and Health” [54] briefly discussed older adults’ sexual health and rights. These reports emphasise the importance of providing information on sexual health to older adults, recognise a need for sexual health services directed towards older adults and call for the elimination of all forms of elder abuse and neglect. However, in none of the reports are older adults mentioned as a potential risk group for SV.

### Health care workers’ response to sexual health needs of older adults

Although policy makers increasingly recognise the importance of sexual health in older age, in practice sexual health needs of older adults remain mostly unmet. A recent European study found that only 6.8% of older women and 12.1% of older men had sought professional help for a sexual difficulty in the past 5 years, and of those who sought help, only 48% were satisfied or very satisfied with the help received [55]. The primary care physician was identified as the main source of professional help [55]. Another study identified the main barrier for older adults to seek help for sexual problems as ‘thinking of sexual changes as normal with age’. Older adults were more likely to seek help if the doctor had asked about sexual function during a routine visit in the last 3 years [56]. Yet, research showed that both primary care physicians and nurses working in long term care tend not to address sexual health proactively when with older adults, as they feel it is not a legitimate topic to discuss with this age group and are afraid to offend their patients [57, 58]. Additionally, primary care physicians tend to look at sexual health in older age in the context of dysfunctions or diseases [59] and consequently discuss sexuality mostly in conjunction with other medical conditions [60].

It seems health care workers, although theoretically recognising that sexuality plays an important role in the lives of older adults, are responding to stereotypes of the ‘asexual older adult’ [57, 58], an image that is often portrayed in the media and has become a wide societal image [61-63].

In Québec, Canada, a group of scientists, practitioners and policy makers revised the current definition of elder abuse and neglect. They included “sexual neglect” into the definition which they define as “a failure to provide privacy, failure to respect a person’s sexual orientation or gender identity, treating older adults as asexual beings and/or preventing them from expressing their sexuality, etc.” [64]. And so, treating older adults as asexual beings could in itself be seen as an act of violence.

## DISCUSSION

Our review showed that 0% to 3.1% of older adults in Europe was sexually victimised in the last 12 months. Lifetime prevalence of SV was 6.3%, which is low knowing that 30% of European women and 10-27% of men experienced at least one incident of SV between the age of 16 and 25 [5-11].

However, the prevalence of SV at older age is likely to be underestimated because of several methodological shortcomings in the existing literature. First, most studies only include questions on penetrative forms of SV like rape and attempted rape, which are less common than for example sexual harassment or sexual abuse without penetration. Second, all studies use data collection methods in which the anonymity of victims is not guaranteed, conceivably leading to safety issues especially when victim and assailant live together. Third, almost all studies excluded cognitively impaired older adults who are known to be at higher risk for several types of abuse [23]. Fourth, older adults may have a different perception of SV than younger generations. In most European countries marital rape only became a criminal offence at the end of the 20^th^ century. Differences in views on SV within intimate partner relationships between older adults and younger generations, may lead to fewer disclosures of SV at older age. Finally, all studies focus on assailants known to the victim, excluding situations where older adults are sexually offended by strangers.

Furthermore, SV in older adults remains embedded in, and consequently diluted by, the broader context of elder abuse and neglect. Detailed reports of SV in older adults are impaired by the wider focus on different forms of abuse (i.e. psychological, physical, financial, and neglect) and an overrepresentation of psychological and financial abuse compared to sexual abuse. In addition, the focus on elder abuse and neglect ignores the fact that older adults may continue to cope with the consequences of SV victimisation in early life, although several studies suggest an important health impact of CSA in later life [18-21]. Also, further research on vulnerability factors for SV in older adults is needed because none of the 14 studies reported on these factors.

Despite our findings, and those of previous research, showing that sexuality remains important in older age [26, 27], older adults are often considered “asexual” in policies and practices [57, 63], leading to an inadequate or even non-existent response to sexual health issues in older age [55, 57]. This assumption of asexuality may further mask the occurrence of sexual victimisation of older adults and their need for tailored care. It is known that SV can induce long-lasting sexual, physical and mental health problems [15]. However, in older adults, manifestations of these consequences are rarely recognized or linked to sexual victimisation [24].

Although leading organizations in the field of health care policy, including the WHO, are starting to recognise the importance of sexual health at older age, they do not acknowledge the complexity of SV in older adults. For example, the WHO’s current definition of elder abuse and neglect does not encompass “sexual neglect”. This omission could lead to the under recognition and inadequate care of older victims of SV. Including this concept in the definition of elder abuse and neglect is important [64]. Adopting a holistic definition in future research and policies could yield more realistic figures on the magnitude and nature of SV in later life, leading to better and more tailored care for future victims and the development of preventive measures for the general public.

## CONCLUSION

Current research suggests that SV in older adults rarely occurs. However, prevalence rates are likely underestimated because of several methodological shortcomings. Ongoing research cannot fully grasp the complexity of SV in older adults by conflating it with other types of violence in the broader context of elder abuse and neglect and by ignoring the possibility of earlier experiences with SV having an influence in later life. We recommend future research to investigate SV in older adults independent of other forms of abuse. Such research should be guided by a broad definition of SV that acknowledges the sexual neglect and should apply a life course approach in order to obtain a better understanding of the magnitude, nature and impact of SV on older adults. The findings of this review indicate that the knowledge about SV in older adults is still limited. A greater awareness about this topic by researchers, policy makers, and health care professionals could contribute to a revision of current policies and health care practices, leading to preventive measures and more tailored care for older victims of SV.

## Data Availability

There was no dataset used in this manuscript

